# Intracranial Directed Connectivity Links Subregions of the Prefrontal Cortex to Major Depression

**DOI:** 10.1101/2024.08.07.24311546

**Authors:** John Myers, Jiayang Xiao, Raissa Mathura, Ben Shofty, Victoria Pirtle, Joshua Adkinson, Anusha B. Allawala, Adrish Anand, Ron Gadot, Ricardo Najera, Hernan G. Rey, Sanjay J. Mathew, Kelly Bijanki, Garrett Banks, Andrew Watrous, Eleonora Bartoli, Sarah R. Heilbronner, Nicole Provenza, Wayne K. Goodman, Nader Pouratian, Benjamin Y. Hayden, Sameer A. Sheth

**Affiliations:** Baylor College of Medicine, Department of Neurosurgery; Brown University, Department of Biomedical Engineering; Baylor College of Medicine, Department of Psychiatry and Behavioral Science; University of Texas: Southwestern, Department of Neurological Surgery

**Author notes:** <u>Corresponding author</u> John Myers, Ph.D., Baylor College of Medicine Department of Neurosurgery Houston, TX 77030.

**Keywords:** depression, prefrontal cortex, directed connectivity, Granger causality

## Abstract

Understanding the neural basis of major depressive disorder (MDD) is vital to guiding neuromodulatory treatments. The available evidence supports the hypothesis that MDD is fundamentally a disease of cortical disinhibition, where breakdowns of inhibitory neural systems lead to diminished emotion regulation and intrusive ruminations. Recent research also points towards network changes in the brain, especially within the prefrontal cortex (PFC), as primary sources of MDD etiology. However, due to limitations in spatiotemporal resolution and clinical opportunities for intracranial recordings, this hypothesis has not been directly tested. We recorded intracranial EEG from the dorsolateral (dlPFC), orbitofrontal (OFC), and anterior cingulate cortices (ACC) in neurosurgical patients with MDD. We measured daily fluctuations in self-reported depression severity alongside directed connectivity between these PFC subregions. We focused primarily on delta oscillations (1-3 Hz), which have been linked to GABAergic inhibitory control and intracortical communication. Depression symptoms worsened when connectivity within the left vs. right PFC became imbalanced. In the left hemisphere, all directed connectivity towards the ACC, from the dlPFC and OFC, was positively correlated with depression severity. In the right hemisphere, directed connectivity between the OFC and dlPFC increased with depression severity as well. This is the first evidence that delta oscillations flowing between prefrontal subregions transiently increase intensity when people are experiencing more negative mood. These findings support the overarching hypothesis that MDD worsens with prefrontal disinhibition.

## Introduction

Major depressive disorder (MDD) is perhaps the most devasting of all mood disorders, labeled by the World Health Organization as the single largest contributor to global disability (Lyus, Buamah, Pollock, Cosgrove, L., & Brhlikova, 2023; WHO, 2017). Although clinicians and researchers have made considerable progress towards treating and understanding MDD, the neural basis of the disorder remains perplexing (Mayberg et al., 2005; Xiao et al., 2023). Depression severity varies across time, but the brain activity associated with these changes has received only modest attention within psychiatry (Fried et al., 2016; Johnson et al., 2016; Xiao et al., 2023). Even within a 24-hour period, fluctuations in depression severity can be used to evaluate treatment effects (Johnson et al., 2016). Studying the neural basis of MDD while accounting for its temporal dynamics has become more crucial as neuromodulatory therapies are further explored (Holtzheimer et al., 2017; Sheth et al., 2021). Some longitudinal work has focused on structural changes to brain structures, highlighting increased brainstem white matter volume as a biomarker for MDD (Soriano-mas et al., 2010). Others have used neuroimaging to measure functional connectivity between the amygdala and dorsolateral prefrontal cortex (dlPFC), reporting that depression severity correlated with decreased prefrontal-limbic connectivity over time (Sacchet, Tymo, Simmons, & Yang, 2017). Most research in this domain has been longitudinal, following participants for several months or years, and they often relied on indirect, noninvasive measurements of neural activity.

Few studies have focused on how the electrophysiological activity underlying depression severity, or even mood in general, varies across hours or days (Sani et al., 2018; Xiao et al., 2023). Sani and colleagues (2018) combined intracranial EEG recordings from the orbitofrontal cortex (OFC), anterior cingulate cortex (ACC), insula, amygdala, and hippocampus with machine learning techniques to decode continuous mood state. Spectral power within the OFC was found to be the most common distinguishing feature of mood shifts, but the study did not focus directly on major depression (Sani et al., 2018). Similarly, Xiao and colleagues (2023) reported that depression severity was correlated with increased low frequency activity and decreased high frequency activity in frontotemporal regions. Although these studies have made impressive strides towards understanding fluctuations in human mood and major depression, neither study focused on connectivity, despite the fact that MDD is well-established as a brain network/system disorder (Kaiser, Andrews-Hanna, Wager, & Pizzagalli, 2015; Linkenkaer-Hansen et al., 2005; Northoff, Wiebking, Feinberg, & Panksepp, 2011). Without understanding which changes in connectivity correspond to dynamic shifts in depression severity, neuromodulation targeting PFC subregions cannot be optimized, and therapy for MDD cannot be improved.

Widespread dysfunction throughout the limbic system and neocortex is associated with depression severity (Drysdale et al., 2017; Fingelkurts & Fingelkurts, 2015; Kaiser et al., 2015; Rolls et al., 2019). The brain regions comprising the limbic system, including the amygdala, ventral striatum, and anterior cingulate cortex (ACC), show altered functional connectivity compared to controls (Drysdale et al., 2017; Rolls et al., 2019). Patients with MDD often have hyperconnectivity within the default mode network and hypoconnectivity within the frontoparietal dorsal attention network (Kaiser et al., 2015; Tozzi et al., 2024). Compared to healthy controls, MDD patients also have hypoconnectivity between the right dlPFC and the right ACC (Y. Wang, Yang, Sun, Shi, & Duan, 2016). Although neuroimaging studies can provide only an indirect glimpse at neuronal communication, there is considerable evidence that maladaptive patterns of connectivity are the underlying causes of the intrusive ruminations associated with depression (B. J. Li et al., 2018; Mogg & Bradley, 2005; Wenzlaff, Wegner, & Roper, 1988). The critical importance of PFC subregions in MDD has become clearer over time (Howard et al., 2019; Pizzagalli & Roberts, 2022), but it is unknown how electrophysiological communication *between* PFC subregions is related to depression severity.

Cognitive processes such as attention and emotion regulation are disrupted in MDD (Loeffler et al., 2018; Rolls, Cheng, & Feng, 2020; L. Wang et al., 2008; Zheng et al., 2018). Subregions of the PFC play distinct roles in these processes (Dixon, Thiruchselvam, Todd, & Christoff, 2017; Pizzagalli & Roberts, 2022; Rudebeck & Murray, 2014). The dlPFC is well known for its dual role in executive function and working memory, which facilitates attention control and prevents emotional distractions (Fales et al., 2008; Heller et al., 2013; Matsuo et al., 2007; L. Wang et al., 2008). The dlPFC also coactivates with limbic structures, which further implicates the region as important for affective processing (Jung, Lambon Ralph, & Jackson, 2022). Historically, the orbitofrontal cortex (OFC) is thought to play a causal role in mood and emotion regulation (Harlow, 1848; Schoenbaum, Setlow, Nugent, Saddoris, & Gallagher, 2003; Szczepanski & Knight, 2014). Electrical stimulation to the OFC can cause dose-dependent improvements in mood (Rao et al., 2018). In cases of brain injury, patients with OFC lesions have diminished responses to emotionally salient stimuli, poor behavioral adaptation, and an increased likelihood of developing MDD symptoms (Drevets, 2007; MacFall, J. R., Payne, Provenzale, & Krishnan, 2001). Compared to healthy controls, the OFC has shown hypoconnectivity with the ACC in MDD patients, but hyperconnectivity with the right dlPFC (Frodl et al., 2010). During working memory tasks, hypoconnectivity within the fronto-parietal network is also characteristic of MDD patients (Vasic, Walter, Sambataro, & Wolf, 2009). Across the literature as a whole, activity patterns within the OFC, dlPFC, and ACC have been highlighted as predictive biomarkers for MDD treatment effects (Lai, 2021). Thus, given the above considerations, we focused on directed connectivity between the OFC, dlPFC, and the ACC in this study.

Much of the research linking brain network connectivity to MDD has used functional magnetic resonance imaging (fMRI) as the sole measure of neural activity (Kaiser et al., 2015; Pizzagalli & Roberts, 2022; Vasic et al., 2009; Y. Wang et al., 2016). However, drawing causal inferences from fMRI data generates a unique set of challenges (Ramsey et al., 2010). Given the high correlations between blood-oxygen level-dependent (BOLD) signals and gamma oscillations (> 50 Hz), interpreting neuronal connectivity can be problematic (Carmichael et al., 2024; Ramsey et al., 2010). The brain is well known to communicate electrochemically (Bucher & Wightman, 2015), and longer-range, even intralaminar, connectivity is more associated with low frequency oscillations (< 15 Hz) (Carracedo et al., 2013; Myers et al., 2022). Low frequency oscillations can even be anticorrelated with the BOLD signal (Feige et al., 2005), which suggests that brain metabolism could *decrease* with widespread cortical synchronization (Pang & Robinson, 2018). Notably, at least one study reported that low frequency oscillations *increased* with the BOLD signal, albeit in anesthetized rats (H. Lu et al., 2007). Aside from the aforementioned link to gamma oscillations, the relationship between the BOLD signal and neural oscillations remains inconsistent (Carmichael et al., 2024; Logothetis, 2003). Therefore, for the purpose of interpreting our results, we hold previous work relevant to the *electrophysiology* of MDD in higher regard.

Parallel to the neuroimaging work on MDD, several lines of research have focused more specifically on the electrophysiological, molecular, and transcriptomic bases of MDD (Duman, Sanacora, & Krystal, 2019; Howard et al., 2019; Pizzagalli & Roberts, 2022). There is strong evidence that the cerebral atrophy associated with MDD is linked to stress-induced excitotoxicity and inflammation (Popoli, Yan, McEwen, & Sanacora, 2013; Wray et al., 2018). The prefrontal ‘hypoactivation’ (compared to healthy controls) observed in neuroimaging studies may be due to excitotoxicity driven by increased intracellular metabolism and oxidative stress (Y. R. Lu, Pei, Xie, & Li, 2024). The major inhibitory neurotransmitter system within the brain, gamma aminobutyric acid (GABA), is compromised in MDD (Fee, Banasr, & Sibille, 2017). This breakdown of inhibitory control is likely preventing the healthy modulation of excitatory neurotransmission for MDD patients (Ghosal, Hare, & Duman, 2017). The two major ‘families’ of GABAergic neurons, parvalbumin-positive and somatostatin-positive neurons, are differentially linked to delta oscillations (∼1-3 Hz) (Cardin, 2019; Kuki et al., 2015), and *both* are damaged in MDD (Duman et al., 2019). Mouse models suggest that delta power *decreases* when knocking out parvalbumin-positive neurons, but *increases* when knocking out somatostatin-positive neurons (Kuki et al., 2015). Somatostatin-positive GABAergic neurons innervate neuronal dendrites (London & Häusser, 2005), and they play a crucial role in synaptic input integration (Song, Yoon, & Lee, 2021). Thus, delta oscillations may be partially driven by the influence of somatostatin-positive neurons (Kuki et al., 2015; Zielinski et al., 2019).

Classical theories on the origins of low frequency extracellular oscillations suggest that delta reflects the summation of enduring after-hyperpolarizations within pyramidal neurons of layer V, rather than synaptic activity per se (G. Buzsaki et al., 1988; Buzsaki, 2002). Although there is heterogeneity within Layer V, a substantial portion of pyramidal cells within are cortical output cells (Brown & Hestrin, 2009; Jones, 2000). ‘Down states’ of these pyramidal cells were thought to produce the extracellular delta oscillations observed in electrophysiology (Buzsáki, Anastassiou, & Koch, 2012; Sirota & Buzsáki, 2005). Although there remains considerable debate about the meaning of low frequency oscillations, their precise phase has been causally linked to cognitive performance measures such as reaction time (Stefanics et al., 2010), and the rhythmic timing of sensory selection (Schroeder & Lakatos, 2009). Delta oscillations have also been linked to many neurological and neuropsychiatric disorders, ranging from sleep disorders (Massicotte-Marquez et al., 2005) to depression (Massicotte-Marquez et al., 2005; Nelson et al., 2018). Delta is prominent in the frontal cortex (Harmony, 2013), and plays an important role in wakeful resting state (H. Lu et al., 2007) and sleep (Lanquart, Nardone, Hubain, Loas, & Linkowski, 2018). Delta power even increases during both physical pain (Bromm, Meier, & Scharein, 1989; Y. D. Li et al., 2020) and panic attacks (Lesser, Poland, Holcomb, & Rose, 1985), which supports the theory that delta oscillations are linked to interoceptive monitoring as well (Knyazev, 2012). These considerations underscore the potential for resting state delta oscillations within the PFC as key biomarkers that signify neuronal communication underlying negative mood and disrupted emotion regulation.

We hypothesized that delta band directed connectivity, measured by Granger causality between PFC subregions, would reveal the directional patterns of activity underlying transient shifts in mood (i.e., higher vs. lower depression severity). As part of a clinical trial utilizing DBS to alleviate treatment-resistant depression (TRD) (UH3 NS103549), we implanted six patients with intracranial stereo EEG (sEEG) electrodes and recorded neural oscillations within the dlPFC, OFC, and ACC (Figure 1A). We found that directed connectivity across these PFC subregions increased with depression severity. Each hemisphere of the PFC also showed distinct patterns of directed connectivity, where 1) strong biases involving connectivity with the left ACC, and 2) increased communication between the OFC and dlPFC in the right hemisphere, were both linked to transient increases in depression severity. The results of this study provide evidence that the delta oscillations flowing across the PFC increase their intensity alongside negative shifts in mood.

**Figure 1.**
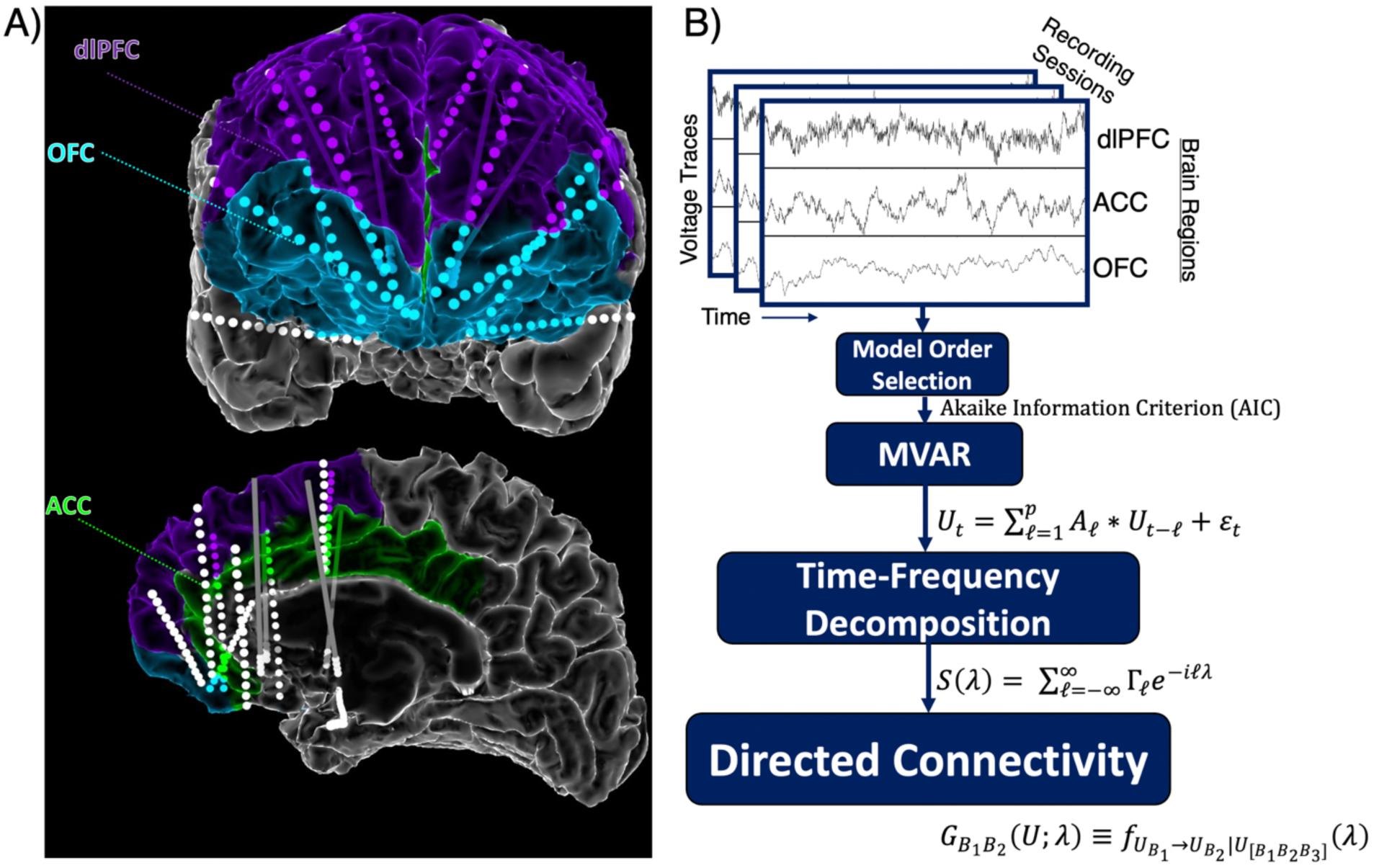
Intracranial EEG and directed connectivity. **A)** All participants had intracranial electrodes bilaterally implanted in the prefrontal cortex (PFC). The prefrontal subregions of interest were the dorsolateral prefrontal cortex (dlPFC), the orbitofrontal cortex (OFC), and the anterior cingulate cortex (ACC). **B)** Analysis pipeline shows progression from raw intracranial EEG data to directed connectivity using MVAR models (see Methods).

## Results

### Depression severity increases with causal connectivity across the PFC

We recorded iEEG from 6 neurosurgical patients (3 male, 3 female) undergoing deep brain stimulation for TRD. Temporary iEEG electrodes were implanted in the dlPFC (Area 10/46), OFC (Area 11), and ACC (Area 24/32) (Figure 1). Throughout a ∼10-day inpatient monitoring period, we acquired n = 78 assessments of depression severity using the computerized adaptive depression inventory (CAT-DI; 13.0 assessments per patient, std.error = 1.97). CAT-DI scores were approximately normally distributed, ranging from ‘normal’ to ‘severe’ (27.0-86.0), and varied similarly across time within subjects (see Methods). Resting state iEEG data was recorded as participants focused their eyes on a fixation cross at center-screen on a computer monitor for 5-minutes duration. For all participants, we selected single grey matter contacts for each PFC subregion (i.e., dlPFC, OFC, ACC), and we measured directed connectivity between them via Granger causality (G-causality: see Methods). Our primary goal was to determine how directed connectivity between PFC subregions is correlated with depression severity.

Directed connectivity from the left OFC to the left dlPFC increased with depression severity, *z*(74) = 3.82, *p* < 0.001). Similarly, directed connectivity from the left dlPFC to the left OFC was also positively correlated with depression severity, *z*(74) = 2.27, *p* = 0.035) (Figure 2, 3A). The statistically significant interaction between these two pathways (i.e., *OFC_left_* → *dlPFC_left_* ∗ *dlPFC_left_* → *OFC_left_*) suggests that *bidirectional communication* between the left OFC and left dlPFC is directly linked to transient shifts in mood, *z*(74) = -4.97, *p* < 0.001). Depression severity was highest when *both* regions exchanged information (Figure 3A). These effects were nearly mirrored in the right hemisphere, but directed connectivity from the right dlPFC to right OFC only marginally increased with depression severity, *z*(74) = 1.92, *p* = 0.055.

**Figure 2.**
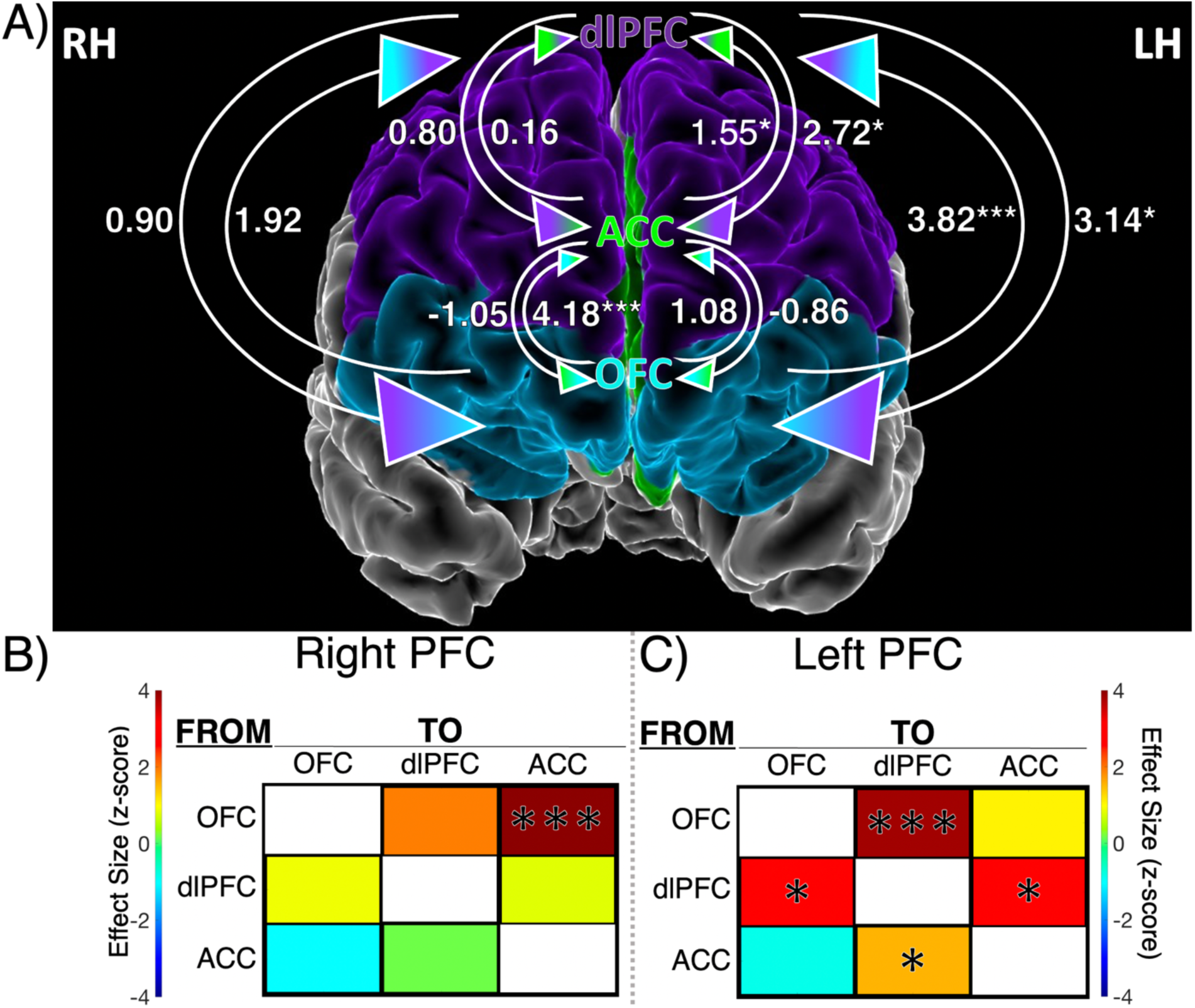
Directed connectivity pathways between PFC subregions are differentially linked to depression severity. **A)** Arrows indicate the direction of connectivity between PFC subregions. The numbers are model coefficients (z-scores) from generalized estimating equations (GEEs) showing the relationship between each individual pathway and depression severity (see Methods). Colormaps show graphical representations of the model coefficients in the right PFC **(B)** and left PFC **(C)**. Stars indicate statistical significance, \**p* < 0.05, \*\*\**p* < 0.001.

**Figure 3.**
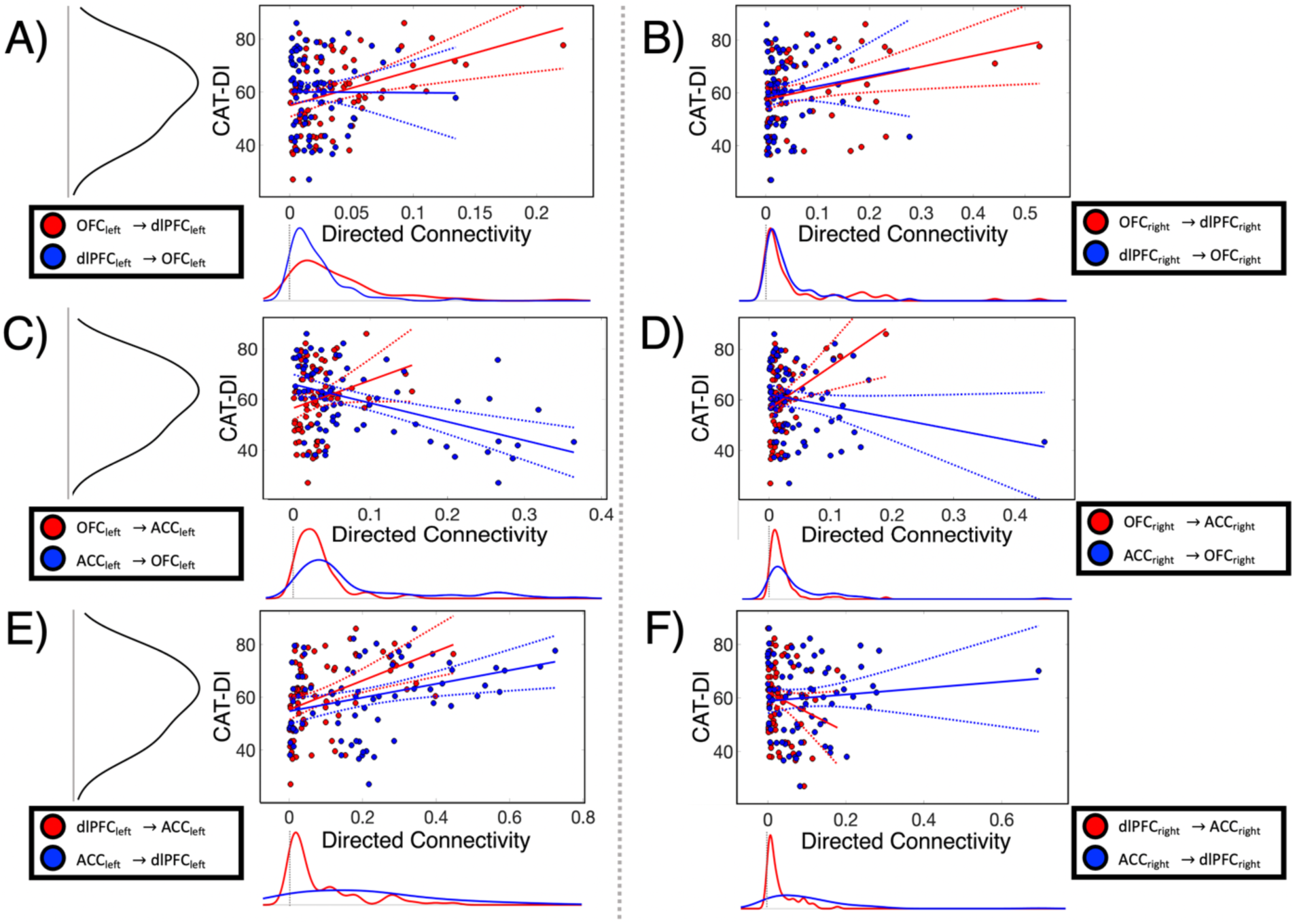
A closer look at interactions between PFC subregions and their relation to depression severity. Scatter plots show the relationships between directed connectivity and depression severity. Depression severity was measured via CAT-DI (y-axes). The x-axes indicate directed connectivity between the subregions labeled subregions in the legends. Significant effects on depression severity were observed in **A**, **D,** and **E**, each with comparable but distinct relationships to depression (*p* < 0.001). Consistently, directed connectivity *from* the OFC to the ACC was positively correlated with depression severity (*p* < 0.001) **(C-D)**. The bidirectional interaction between the right OFC and ACC was marginally significant (*p* = 0.055) (**D**). These findings expand our understanding on the role of PFC subregions in depression. Regression lines were fit to the data using robust (bi-square) regression for visualization purposes. Dotted lines indicate confidence bounds. Kernel density plots on the x- and y-axes display probability distributions of the corresponding variables.

Bidirectional communication between the left dlPFC and left ACC also increased with depression severity, as indicated by the significant *dlPFC_left_* → *ACC_left_* ∗ *ACC_left_* → *dlPFC_left_* interaction, *z*(74) = -2.32, *p* = 0.020 (Figure 3E). Directed connectivity from the left dlPFC to the left ACC was positively correlated with depression severity, [*dlPFC_left_* → *ACC_left_*; *z*(74) = 2.66, *p* = 0.008], as was its counterpart *ACC_left_* → *dlPFC_left_*; *z*(74) = 2.31, *p* = 0.020. In the left hemisphere, bidirectional communication between the OFC and ACC was not significantly linked to depression severity, *z*(74) = - 1.18, *p* = 0.158 (Figure 2A,C, 3C). In the right hemisphere, we observed a marginally significant *OFC_right_* → *ACC_right_* ∗ *ACC_right_* → *OFC_right_* interaction, which may indicate that bidirectional communication between the right OFC and right ACC had a more pronounced effect on mood compared to the left hemispheric counterparts, *z*(74) = - 1.92, *p* = 0.055. Indeed, directed connectivity from the right OFC to the right ACC significantly increased with depression severity, [*z*(74) = 4.18, *p* < 0.001], whereas the left hemisphere homologue showed no effect, *z*(74) = 1.08, *p* = 0.279 (Figure 2, 3C-D).

The strongest initial distinction between hemispheres was the greater number of significant main effects in the left PFC (4 main effects) vs. the right PFC (1 main effect) (Figure 2B,C). However, a t-test comparing left PFC vs. right PFC effect sizes (i.e., coefficients from generalized estimating equations (GEEs)) showed no significant difference between hemispheres in their relation to depression severity, *t*(5) = 0.92, *p* = 0.402. In accordance with this quantitative comparison, the pattern of directed connectivity effects was also similar across hemispheres (Figure 2B,C) and individual participants (Figure S1). Taken together, these results indicate that PFC subregions increase their communication alongside transient increases in depression severity. Increased intra-PFC communication may indicate negative shifts in mood and increased attention to emotion regulation.

### Comparative strength of communication within each PFC hemisphere is directly linked to depression severity

A cursory interpretation of the above analyses might suggest that both hemispheres of the PFC play statistically equivalent roles in depression. However, a series of more direct comparisons between PFC hemispheres revealed stark differences in their relation to depression severity. We first conducted pairwise comparisons between the left PFC and right PFC homologues (e.g., *ACC_left_* → *dlPFC_left_ versus ACC_right_* → *dlPFC_right_*) to determine which hemisphere had stronger inter-regional communication. All communication involving the ACC was higher in the left hemisphere compared to the right hemisphere (Figure 4). Directed connectivity from the ACC to the dlPFC was significantly higher in the left PFC, *t*(77) = 6.20, *p* < 0.001. Similarly, directed connectivity from the left dlPFC to the ACC was also higher in the left PFC, *t*(77) = 4.21, *p* < 0.001. Directed connectivity from the OFC to the ACC, [*t*(77) = 2.52, *p* = 0.014], as well as from the ACC to the OFC, were also higher in the left hemisphere, *t*(77) = 3.06, *p* = 0.005.

**Figure 4.**
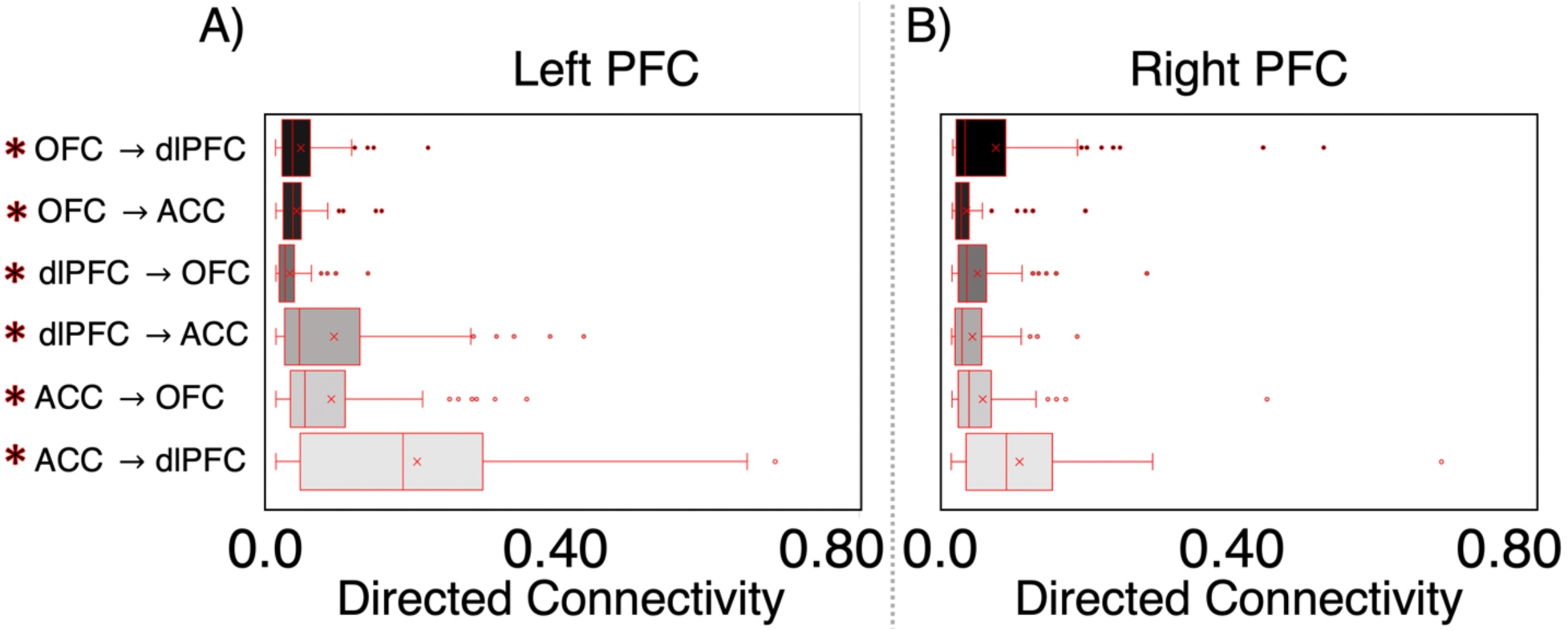
Directed connectivity magnitudes across PFC subregions in each hemisphere. Six paired t-tests were conducted between the left PFC and right PFC homologues to determine which hemisphere had stronger inter-regional communication. All communication involving the ACC was higher in the left hemisphere, including directed connectivity from the ACC to the dlPFC (*p* < 0.001), dlPFC to ACC (*p* < 0.001), OFC to ACC (*p* = 0.014), and from the ACC to the OFC (*p* = 0.005). In the right hemisphere, directed connectivity from the OFC to the dlPFC (*p* = 0.011), and from the dlPFC to the OFC was higher (*p* = 0.003). These differences in connectivity magnitudes suggest that communication between PFC subregions is specialized in each hemisphere. All t-test results were false-discovery rate (FDR) corrected for the multiple comparisons across hemispheres. X’s within each box show the mean directed connectivity, vertical lines indicate medians. Outliers are indicated by single data points (quartile 3-1.5*interquartile range). **\***asterisks indicate significant differences between hemispheres (*p* < 0.05).

In the right hemisphere, communication was stronger between the OFC and dlPFC. Directed connectivity from the right OFC to the right dlPFC was higher than the left homologue, *t*(77) = -2.66, *p* = 0.011. The reverse was also true, where communication from the dlPFC to the OFC was higher in the right hemisphere, *t*(77) = - 3.23, *p* = 0.003 (Figure 4). These significant differences in connectivity magnitudes across hemispheres inform us about how communication between PFC subregions could be specialized in each hemisphere. Critically, all results reported above were false-discovery rate (FDR) corrected for the multiple comparisons across hemispheres.

Given the observed communicative differences within each PFC hemisphere, we investigated further to determine whether depression severity was linked to a ‘hemispheric differential’, which here refers to the arithmetic difference between directed connectivity in the right-minus-left hemisphere (e.g., *ACC_right_* → *dlPFC_right_* − *ACC_left_* → *dlPFC_left_*). We subjected these hemispheric differential scores to the same GEE analyses outlined in the Methods section to quantify their relationship to depression. Depression severity increased with the hemispheric differential between the right and left *OFC* → *dlPFC*, revealing a strong bias towards the right hemisphere, *z*(74) = 3.69, *p* < 0.001. Depression severity was higher when *OFC_right_* → *dlPFC_right_* was greater than *OFC_left_* → *dlPFC_left_* (Figure 5A). On the other hand, communication between the OFC and ACC showed a left hemisphere bias, where depression severity decreased when *OFC_left_* → *ACC_left_* was greater than *OFC_right_* → *ACC_right_* (Figure 5B), *z*(74) = -4.96, *p* < 0.001. Directed connectivity between the left dlPFC and left ACC was also biased towards the left hemisphere, *z*(74) = -2.06, *p* = 0.039. Depression severity increased when *dlPFC_left_* → *ACC_left_* was greater than *dlPFC_right_* → *ACC_right_*.

**Figure 5.**
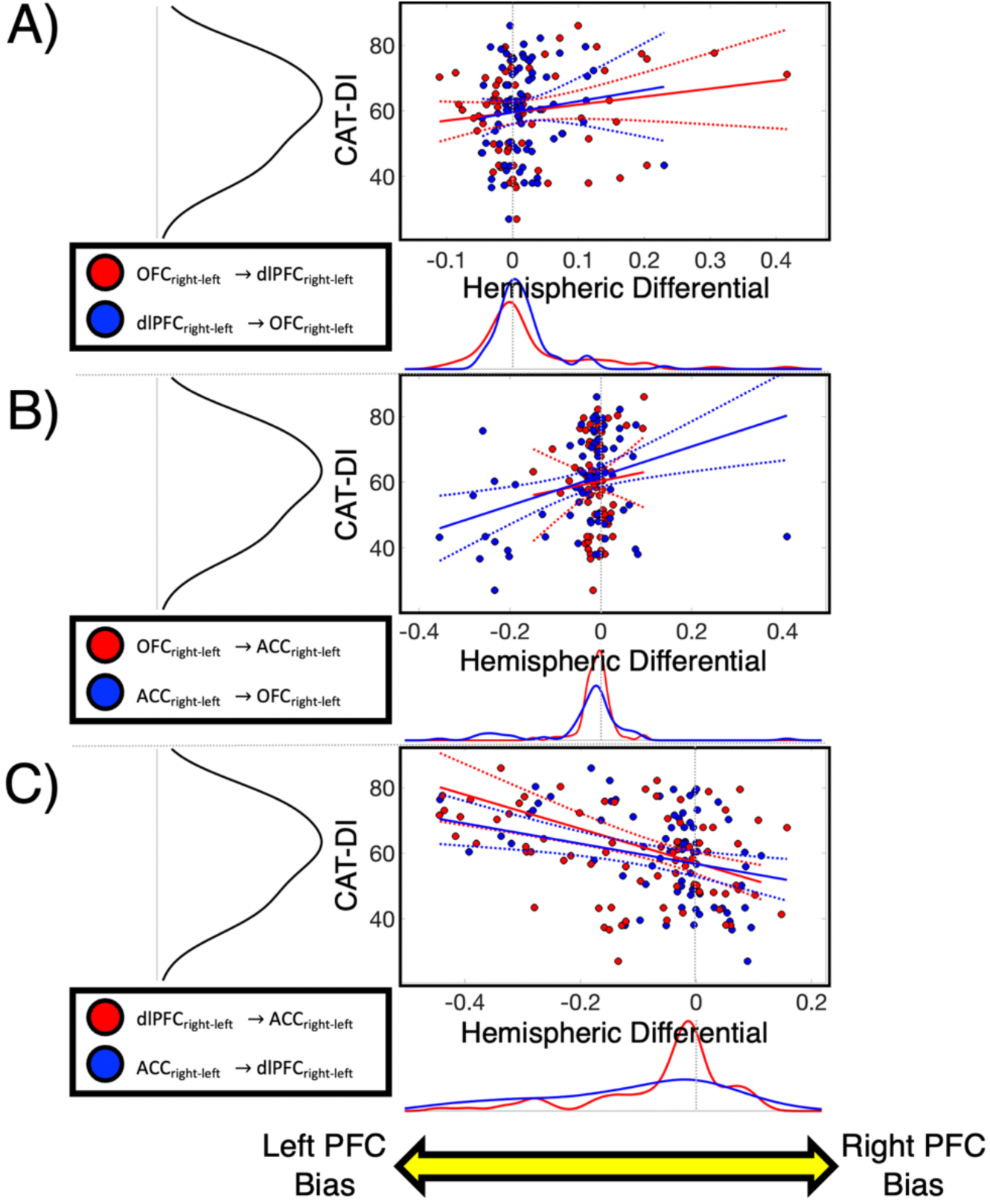
Differences between directed connectivity across hemispheres correlate with depression severity. The ‘hemispheric differential’ (x-axis) refers to the subtractive difference between directed connectivity in the right-minus-left hemisphere. Therefore, more positive differentials indicate a right hemisphere bias, and vice versa for negative differentials (i.e., left bias). In order to quantify their relationship to depression, the hemispheric differential scores were subjected to the same analysis structure depicted in Figure 3. **A)** Depression severity was higher when *OFC_right_* → *dlPFC_right_* was greater than *OFC_left_* → *dlPFC_left_*, which indicates a right hemispheric bias (*p* < 0.001). **B)** Directed connectivity between the OFC and ACC showed a left hemispheric bias, where depression severity decreased when *OFC_left_* → *ACC_left_* was higher than *OFC_right_* → *ACC_right_* (*p* < 0.001). **C)** Communication between the left dlPFC and ACC was also biased towards the left hemisphere (*p* = 0.039). Depression severity increased when *dlPFC_left_* → *ACC_left_* was greater than *dlPFC_right_* → *ACC_right_*. These results provide clear evidence that the right PFC and left PFC play separate roles in major depression.

Collectively, these findings provide clear evidence that the left and right PFC play distinct roles in major depression. The observed differences in directed connectivity magnitudes across hemispheres largely aligned with how the hemispheric differentials correlated with depression severity. The hemispheric differential results further support the evidence that directed connectivity involving the left ACC is closely linked to depression severity (Figure 5A,C). However, in the right hemisphere, communication between the OFC and dlPFC proved to be more linked to depression severity than the same regions within the left hemisphere (Figure 5A).

### Delta spectral power within PFC subregions is differentially linked to depression severity

Although our primary focus in this work involved prefrontal connectivity, we also measured power spectral density (PSD) within the same delta band (1-3 Hz) to determine how local energy within PFC subregions, instead of communication between them, was related to depression severity. In the left hemisphere, we observed no effects of delta spectral power on depression severity (Figure 6 A,C,E). However, in the right hemisphere, spectral power within the OFC [*z*(71) = 2.90, *p* = 0.004] and dlPFC were positively correlated with depression severity, *z*(71) = 2.01, *p* = 0.045 (Figure 6 B,D). The significant interaction between the spectra of the right OFC and right dlPFC aligns with the effects we found via directed connectivity, where increased delta power in both regions was positively correlated with depression severity, *z*(71) = 2.23, *p* = 0.025.

**Figure 6.**
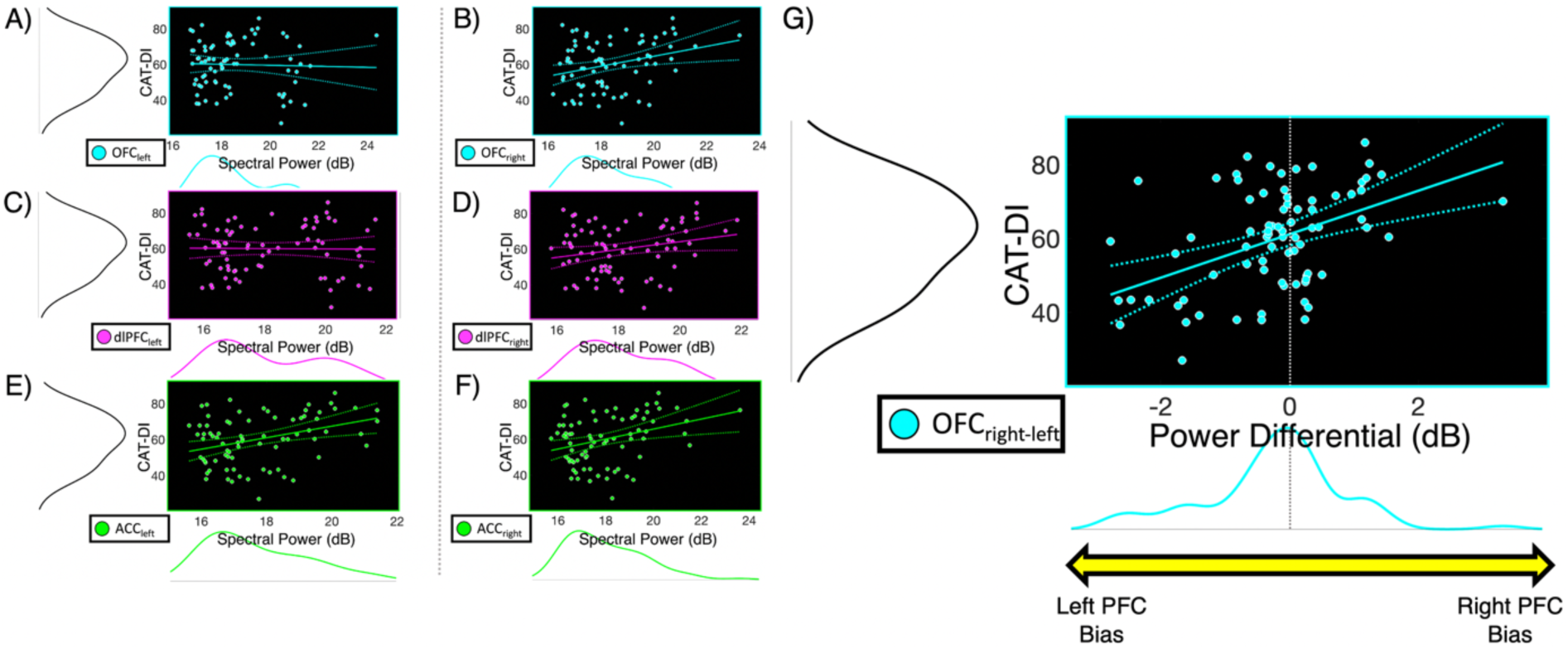
Power spectral density in right PFC subregions is linked to depression severity. **A,C,E**) In the left hemisphere, we observed no effects of delta spectral power on depression severity **B,D,F**) In the right hemisphere, spectral power within the OFC (*p* = 0.004) and dlPFC (*p* = 0.045) were positively correlated with depression severity. **G)** The right-minus-left ‘power differential’ revealed a right hemispheric bias within the OFC, where depression severity increased when delta power within the right OFC was greater than the left OFC (*p* = 0.040).

We also conducted pairwise t-tests to determine which PFC subregions had higher energy across hemispheres. Only the ACC showed significant differences between hemispheres, where the right ACC (mean = 17.99 dB, st.dev = 1.59) had consistently greater spectral power than the left ACC (mean = 17.71 dB, st.dev = 1.52), *t*(77) = -5.23, *p* < 0.001 (Figure 6 E, F). Notably, spectral power within the left OFC was somewhat higher than the right OFC, but the significance did not survive FDR correction, *t*(77) = 1.98, *p* = 0.076. To determine whether there were hemispheric biases in the PSD, we conducted a similar analysis involving the right-minus-left ‘power differential’. We observed a right hemispheric bias within the OFC, where depression severity increased when right OFC delta power was greater than the left OFC, *z*(74) = 2.05, *p* = 0.040 (Figure 6G). Overall, these PSD effects aligned with those observed for directed connectivity.

## Discussion

In this study, we found that transient increases in depression severity were associated with increased directed connectivity between PFC subregions. This is the first research of its kind to show how directed connectivity within the PFC is linked to MDD. Much of the previous work has reported ‘hypoactivation’ of the PFC as a biomarker of depression (George, Ketter, & Post, 1994; Y. R. Lu et al., 2024; Pizzagalli & Roberts, 2022; Zhong et al., 2011). Instead, we found that communication between PFC subregions *increased* alongside depression severity. Our results align more closely with the evidence that MDD is related to diminished cortical inhibition (Duman et al., 2019; Hamilton, Chen, Thomason, Schwartz, & Gotlib, 2011). Our focus on slow, delta oscillations (1-3 Hz) in the PFC provided us with a robust theoretical foundation that facilitated both interpretation and analysis. As previously noted, we focused on delta specifically because of its prominence in frontal cortex (Kiss, Hoffmann, & Hajós, 2011), theorized role in mood disorders (Knyazev, 2012), and its links to GABAergic inhibition (Cardin, 2019; Y. D. Li et al., 2020). There remains much to be discovered within other frequency bands, perhaps with specific attention to phase-amplitude coupling across higher and lower frequencies. Given this new evidence on within-PFC connectivity in depression, future studies can focus even more closely on cross-frequency dynamics.

Directed connectivity *from* the OFC to other frontal regions was always positively correlated with depression severity. These effects may be related to the increased need for inhibitory control during negative shifts in mood, given that the ACC is involved in emotional response inhibition (Albert, López-Martín, Tapia, Montoya, & Carretié, 2012), and the dlPFC is involved in the cognitive control of attention (Smith et al., 2019). The OFC’s well known inhibitory control could be compromised in MDD, resulting in ruminations and poor emotion regulation (Balasubramani & Hayden, 2018; Cooney, Joormann, Eugène, Dennis, & Gotlib, 2010; Rudebeck & Murray, 2014). Patients with MDD tend to have lower gray matter volume in the OFC and ACC, which may explain this increased ‘need’ for input from other PFC regions to help stabilize mood (Bijanki, Hodis, Brumm, Harlynn, & McCormick, 2014; Yu et al., 2018). The OFC and ACC are functionally connected to mood (Frodl et al., 2010), but this is the first evidence that intracranial delta oscillations between these regions are linked to depression severity. The crosstalk between the PFC subregions may relate to frequent self-appraisals of mood during MDD episodes and diminished control of emotional states (Dixon et al., 2017; Golkar et al., 2012).

There has long been evidence that the left and right hemispheres of the brain serve distinct functions in affective processing and MDD (Coffey, 1987; Quigg, Broshek, Heidal-Schiltz, Maedgen, & Bertram, 2003). For example, compared to healthy controls, people with MDD often have abnormal perceptual asymmetries during dichotic listening tasks, revealing pathological biases within the brain’s right hemisphere (Bruder, Stewart, & McGrath, 2017). Discoveries such as these led to a heavy emphasis on right-hemispheric activity in research on MDD (Hecht, 2010). In this study, the most distinguishing feature of the right hemisphere was the strong correlation between depression severity and directed connectivity between the right OFC and right dlPFC (i.e., *OFC_right_* → *dlPFC_right_*). This effect aligns with a previous study focused on trait-based differences between MDD patients and healthy controls (Frodl et al., 2010). By contrast, the most distinguishing feature of the left hemisphere was that all communication pathways involving the left ACC were positively correlated with depression severity. Imbalanced connectivity between the left and right PFC also seems to contribute to MDD severity, as evidenced by our analyses comparing hemispheres (Figure 5A,C).

Both sets of findings from directed connectivity and spectral power analyses align to suggest that communication between and within PFC subregions are distinct in their relationships to major depression. Taken together, these findings support the hypothesis that the left PFC is just as essential for mood regulation as the right PFC (Johnstone, Van Reekum, Urry, Kalin, & Davidson, 2007; Ochsner et al., 2004). The evidence provided here suggests that the two hemispheres play distinct but complementary roles in emotion regulation. Future work may focus on inter-hemispheric PFC interactions across the corpus callosum to elucidate even further. Given that the PFC receives input from several limbic regions (Allawala et al., 2023), the PFC activity observed here was very likely influenced by the amygdala, hippocampus, basal ganglia, and thalamic nuclei as well (Mayberg et al., 1999; Zheng et al., 2018). Future research could employ similar techniques to explore larger scale networks and determine how limbic structures provide information to PFC subregions.

Controlled studies using repetitive transcranial magnetic stimulation (rTMS) to target the dlPFC has reported benefits for MDD patients (Daskalakis, Levinson, & Fitzgerald, 2008; Klein et al., 1999). A proposed mechanism for the therapeutic effects of rTMS is connectivity between the dlPFC and the ACC, especially the subgenual region (Fox, Buckner, White, Greicius, & Pascual-Leone, 2012). Given the bidirectional communication we observed between the left dlPFC and left ACC, combined with their well-known anatomical and functional links (Cieslik et al., 2013), our findings were consistent with this proposition. The dlPFC’s well known role in cognitive attention make it a physiologically appropriate candidate when more attention is focused on negative mood (Cieslik et al., 2013; Kohn et al., 2014). Moreover, data from lesion studies suggest that stroke patients with left dlPFC damage are more likely to develop severe MDD symptoms than patients with right dlPFC damage (Grajny et al., 2016). Although high frequency TMS (∼10 Hz) to the left dlPFC is the conventional FDA-approved target for MDD, *bilateral* stimulation is thought to provide the greatest benefit (Berlim, Van Den Eynde, & Daskalakis, 2013; Daskalakis et al., 2008).

A final key distinction setting this study apart is our focus on more rapid fluctuations in depression severity across days, compared to previous studies that focused on metabolic and structural changes across months or years (Heller et al., 2013; Sacchet et al., 2017; Soriano-mas et al., 2010). Although the clinical opportunities for this research are limited, they are nonetheless invaluable. There are very few TRD patients with intracranial electrodes implanted, and thus sample sizes tend to be much lower compared to larger scale studies of MDD using noninvasive methods (Sheth et al., 2021). However, we have a key advantage of being able to record from participants several times, which allowed us to capture more transient variations of neural activity ad depression symptoms. The depression inventory scores were normally distributed and ranged from normal-to-severe across patients, but the treatment-resistant patient population still represents a subset of individuals with MDD. Forthcoming intracranial research will explore a broader range of patients, with a clear focus on comparing neurosurgical patients with MDD to those without. Our work can serve as a guide to uncovering how specific patterns of activity within PFC networks are linked to other mood disorders as well, such as bipolar disorder. Understanding such patterns of connectivity will certainly pave the way for additional therapeutic innovations, with vast implications for brain stimulation and improved mental health for millions of people.

## Methods

### Participants

Six patients with TRD (3 male; 3 female; avg. age 44.33 y/o) who were implanted with therapeutic DBS leads in the ventral capsule/ventral striatum and sub-callosal cingulate were also implanted with temporary sEEG electrodes in the dlPFC (BA 10/46/47), OFC (BA 11), and ACC (BA 24/32) for neural recordings during a 9-day inpatient monitoring period (see Figure 1A). These patients were part of an NIH-funded clinical trial via the Brain Research through Advancing Innovative Neurotechnologies (BRAIN) Initiative (UH3 NS103549) and were the subjects of a previous study about using machine learning to predict depression severity (Xiao et al., 2023). All patients provided written and verbal consent to participate in this study, which was approved by the Institutional Review Board at Baylor College of Medicine.

### Intracranial Recordings and Preprocessing

Neural data were recorded while participants were sitting with their eyes focused on a fixation cross at center-screen on a computer monitor (5-minutes duration). All signals were amplified and recorded via sEEG electrodes at 2 kHz using a Cerebus data acquisition system (Blackrock Microsystems). Signals were bandpass filtered during the recordings at 0.3-500 Hz via Butterworth filter. Single contacts in the dlPFC, OFC, and ACC were selected by convergence of expert ratings, where authors B.S. and S.H. studied the fused preoperative MRI to postoperative CT images in order to determine whether each contact was within gray matter in the regions of interest (dlPFC, OFC, ACC). Electrode coordinates were acquired via reconstruction in Free-Surfer (https://surfer.nmr.mgh.harvard.edu/).

All neural data were visually inspected for the presence of recording artifacts. Channels that were found to be noisy were excluded from further analysis to prevent spread to other channels via re-reference. Each channel was notch filtered to attenuate line noise (60 Hz and harmonics) and then re-referenced against adjacent channels via bipolar referencing.

### Depression Severity Assessment

We acquired measurements of depression severity throughout a 9-11 day inpatient monitoring period using the computerized adaptive test for depression inventory (CAT-DI) (Gibbons et al., 2012), where higher scores denoted higher depression severity. Each CAT-DI survey question was adaptively selected based on a patient’s previous answers. The CAT-DI can be completed quickly (∼12 questions; ∼3-minutes/test), has high test-retest reliability (*r* ≥ 0.90), and its scores are highly correlated with other depression inventories that can take much longer for patients to complete (Gibbons et al., 2012; Xiao et al., 2023). The survey data were comprised of 13.0 (std.error = 1.96) CAT-DI assessments from each patient (78 total assessments). Depression severity scores ranged from 27.0 to 86.0 (i.e., ‘normal’ to ‘severe’), and a Kolmogorov-Smirnov test indicated an approximately normal distribution (*p* = 0.701) (see Figure 3). Throughout the inpatient stay, depression severity varied similarly across patients, with mean (and standard deviation) of 61.8 (9.18). The mean score falls within the ‘mild’ 50-65 range (Gibbons et al., 2012).

### Directed Connectivity Modeling

We measured directed connectivity using multivariate vector autoregressive (MVAR) models to quantify information flow across the prefrontal network, comprised of the dlPFC, OFC, and ACC. MVAR models are used to estimate Granger causality (G-causality) in multivariate systems of time series data (Barrett, Barnett, & Seth, 2010; Bressler, Kumar, & Singer, 2022; Bressler & Seth, 2011; Krumin & Shoham, 2010). G-causality measures the extent to which a process, *X_t_*_=0_ (i.e., ‘X at time zero’) can be better predicted by knowing the prior state of process, *Y_t-ℓ_* (i.e., ‘Y at time lag ℓ’), relative to only knowing *X_t-ℓ_*. Therefore, *Y_t-ℓ_* can only G-cause *X_t_*_=0_ if prediction error *decreases* when knowing *Y_t-ℓ_*, compared when knowing only *X_t-ℓ_*. Logically, G-causality is based on the premises that (1) causes occur *before* effects, and that (2) knowledge of causes *reduces* prediction error (Granger, 1963). We applied this logic to quantify directed connectivity between PFC subregions involved in MDD.

The G-causality measure of interest in this study is the pairwise-conditional spectral causality, 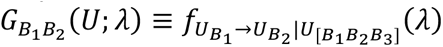, which computes directed connectivity between two brain regions, *B*_1_ → *B*_2_, while controlling for the ‘universe’ of other pairwise relationships, 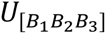 (Barnett & Seth, 2014) (see Figure 1B). MVAR modeling began with a multivariate matrix, *U*_1_, *U*_2_, … containing all brain regions, *B*, for all timepoints, *t*. Each model was constructed as 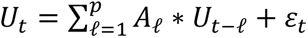, where *p* is the model order determined by minimum Akaike Information Criterion (AIC) from up to 250 milliseconds of data (*model order* ≤ 500) (Akaike, 1974). The regression coefficients, *A*_ℓ_, were estimated using locally weighted regression (LWR) via the Multivariate Granger Causality (MVGC) toolbox in MATLAB (MathWorks) (Barnett & Seth, 2014). All VAR processes were determined to be covariance-stationary by measuring the spectral radius to ensure the functions were invertible within the complex plane (< 1.0) (Lütkepohl, 2005). This stationarity test protects G-causality from the slow, unstable dynamics that could diminish their reliability (Friston et al., 2014). The MVAR autocovariance sequence, Γ_ℓ_ ≡ *cov*(*U_t_*, *U_t-ℓ_*), was Fourier transformed to generate the cross-power spectral density (CPSD) of the process, 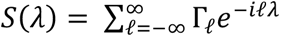. The CSPD was then factorized via transfer function, *H*(*λ*), to *S*(*λ*) = *H*(*λ*) ∗ ∑ *H*(*λ*)^∗^, where *H*(*λ*) is the inverse Fourier transform of the MVAR coefficients. This factorization allows for stable transitions between time and frequency domains, which avoids pitfalls related to separate full vs. reduced model fitting (Barnett, Barrett, & Seth, 2018; Stokes & Purdon, 2017). To take advantage of these frequency domain representations, we focused on G-causality within the delta band (1-3 Hz), given its prominence in the frontal cortex (Harmony, 2013) (Figure S2).

### Power Spectral Density Computations

Power spectral density (PSD) was computed for each PFC contact via wavelet convolution. Each signal, *x*(*t*), from a given electrode, was convolved with a Morlet wavelet defined by 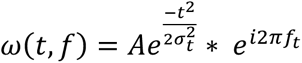, where *A* is the normalized amplitude, such that 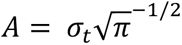 is time, *f* is frequency, and *σ* is wavelet duration. The number of cycles in each wavelet function was 6. All data were analyzed from 1-50 Hz in 0.1 Hz intervals, totaling, 491 frequencies. In alignment with the directed connectivity analyses, we focused on the average PSD from 1-3 Hz.

### Statistical Analyses

We used generalized estimating equations (GEEs) to test how resting state *directed connectivity* (*DC*) correlated with depression severity (*MDD*). The GEEs were constructed as follows. For all subjects *i* = 1,…, *m* at times 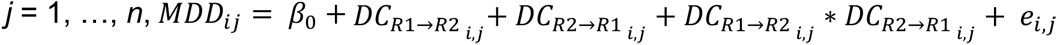. The bidirectional communication terms involving both brain regions, *R1*↔*R2*, were included as well. Given repeated testing of depression severity, these data are inherently autocorrelated within subjects. Thus, estimating the autocorrelation structure was needed to yield proper GEE parameters. The autocorrelation was estimated by 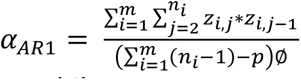, where *z_i, j_* and *z_i, j -1_* were the residuals for subject *i* at time *t_i, j_*, and time *t_i, j -1_*, respectively. The dimension of the intercept is given by *p* and the model-based scaling parameter is ∅ø (Chaganty & Shults, 1999; Ratcliffe & Shults, 2008). GEEs produce data driven estimates of population-level parameters while accounting for correlations within subjects. These models were computed using ‘robust’ fitting within the GEEQBOX (MATLAB; MathWorks) to account for potential outliers (Ratcliffe & Shults, 2008). The advantages of GEE, in contrast to mixed effect models, can provide more accurate estimates of effects within autocorrelated (i.e., repeated measures) data, with fewer statistical assumptions (Chaganty & Shults, 1999; Gardiner, Luo, & Roman, 2009). Each hemisphere of the PFC was analyzed separately to conserve rank for statistical testing and the analysis of interaction effects within hemispheres. There is also evidence that the left and right hemispheres could play distinct roles in MDD (Cook, Hunter, Abrams, Siegman, & Leuchter, 2009; Grajny et al., 2016). In order to test whether depression severity was linked to differences in directed connectivity between hemispheres, we derived a ‘hemispheric differential’, which is the subtractive difference between directed connectivity in the right-minus-left hemisphere (e.g., *OFC_right_* → *dlPFC_right_* − *OFC_left_* → *dlPFC_left_*). The hemispheric and power differential scores were subjected to the same structure of GEE analyses outlined above. All p-values were false discovery rate (FDR) corrected to account for multiple comparisons across hemispheres (Benjamini & Hochberg, 1995). Although our core objective was to measure correlations between G-causality and depression severity, we also tested G-causality against theoretical asymptotic null distributions via Geweke’s chi square test in the MVGC toolbox (Barnett & Seth, 2014; Geweke, 1982). All sessions (n = 78) contained statistically significant Granger causality (i.e., greater than zero, *p* < 0.05).

## Data Availability

All data produced in the present study are available upon reasonable request to the authors

---

**Figure S1.**
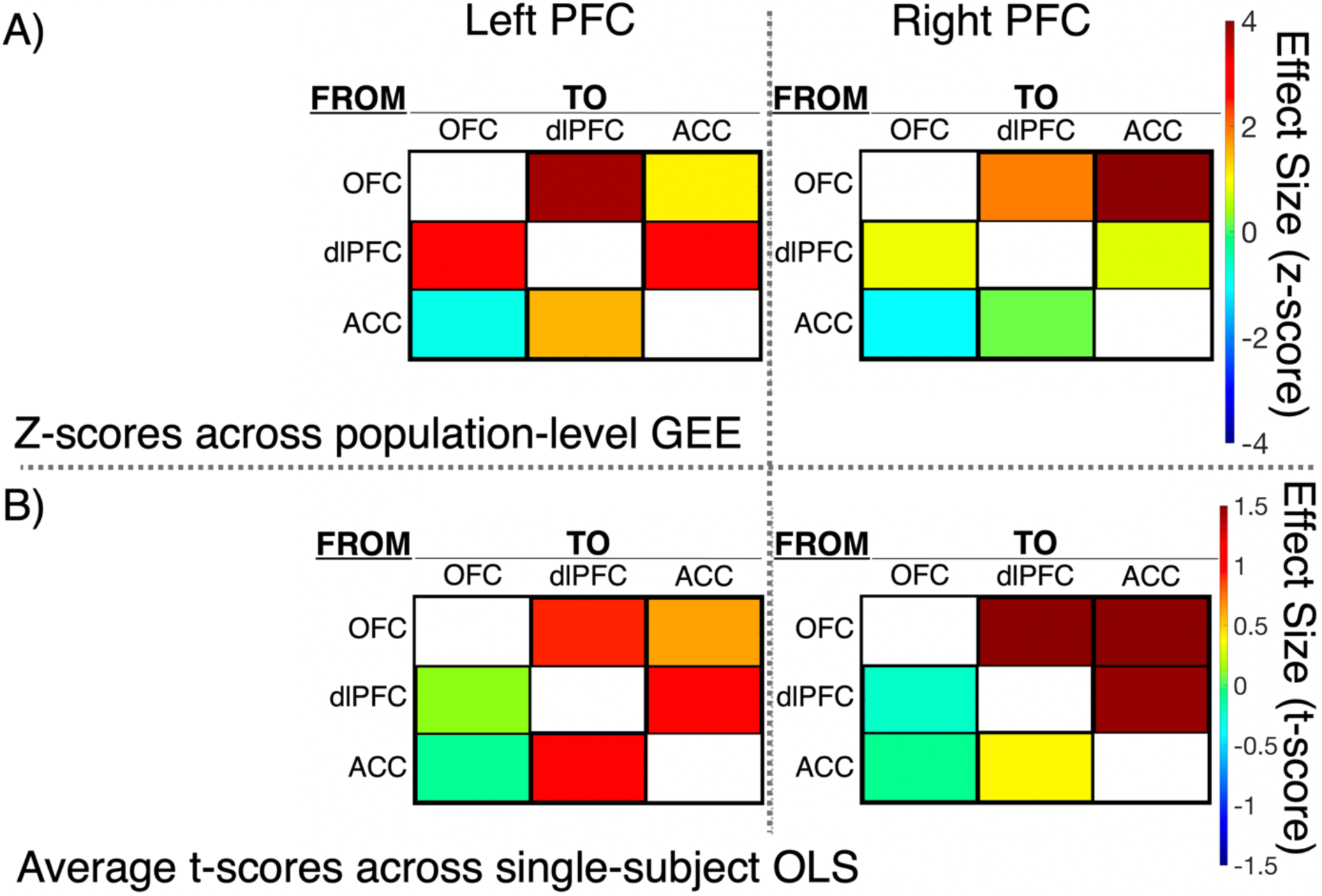
Visual comparisons of population-level results with average single-subject effects. **A)** Colormaps show graphical representations of the population-level GEE coefficients (z-scores) in the left and right PFC **(B)**. Graphical representations of the average single-subject-level ordinary least squares (OLS) coefficients (t-scores). This figure demonstrates how single-subject OLS models averaged across subjects are directionally aligned with the population-level GEE results, which more appropriately took into account variations between patients and autocorrelation within patients across time.

**Figure S2.**
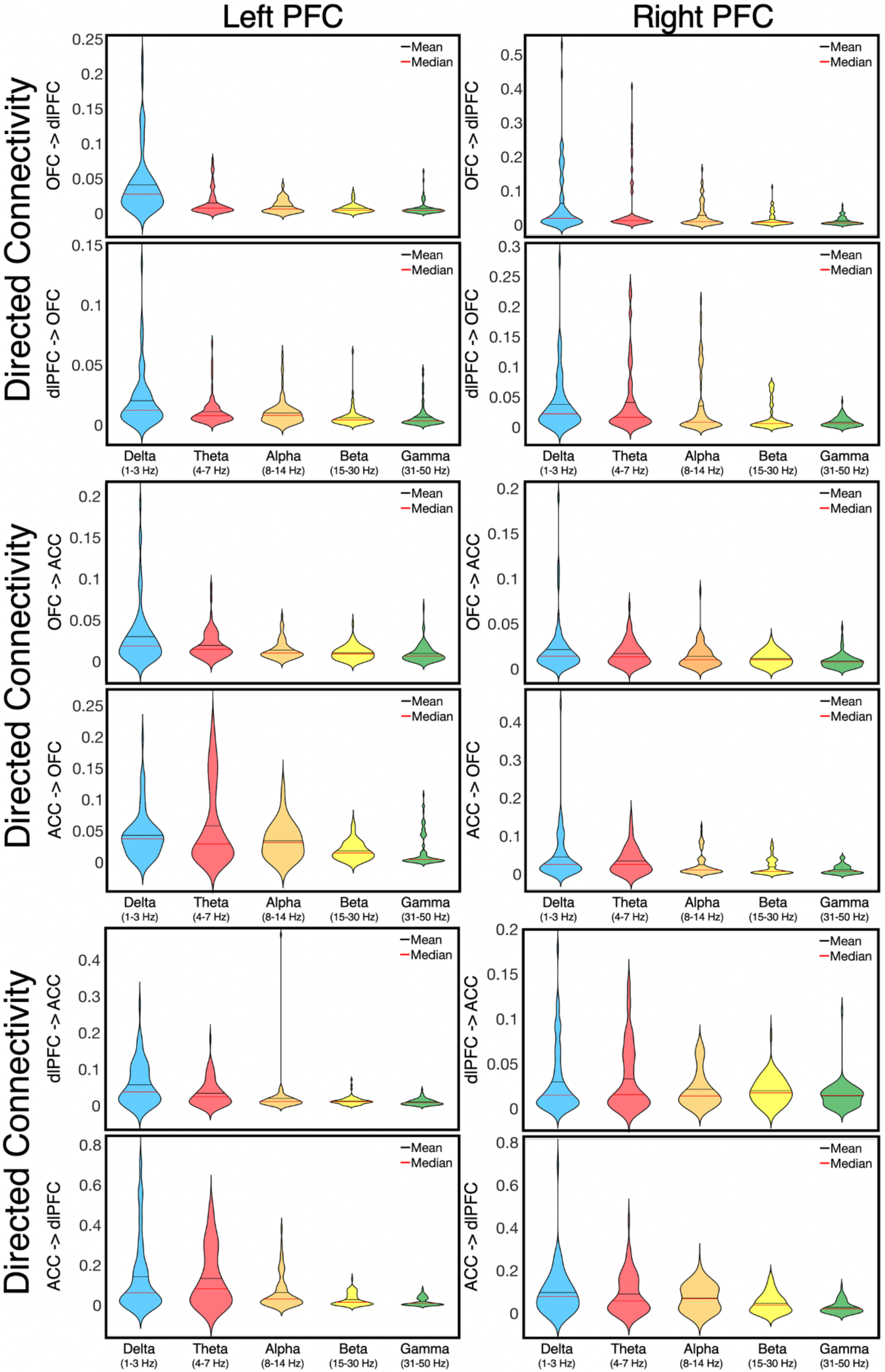
Directed connectivity across frequency bands. **A)** Violin plots show narrowband directed connectivity, including delta (1-3 Hz), theta (4-7 Hz), alpha (8-14 Hz), beta (15-30 Hz), and gamma (31-50 Hz) frequency bands in the left and right PFC. Directed connectivity within the delta band (blue: 1-3 Hz) was the primary focus of this study.

